# Proteomic analysis of circulating immune cells identifies novel cellular phenotypes associated with COVID-19 severity

**DOI:** 10.1101/2022.11.16.22282338

**Authors:** Martin Potts, Alice Fletcher-Etherington, Katie Nightingale, Federica Mescia, Laura Bergamaschi, Fernando J. Calero-Nieto, Robin Antrobus, James Williamson, Cambridge Institute of Therapeutic Immunology and Infectious Disease-National Institute of Health Research (CITIID-NIHR) COVID BioResource Collaboration, Nathalie Kingston, Berthold Göttgens, John R Bradley, Paul J Lehner, Nicholas J Matheson, Kenneth G.C. Smith, Mark R Wills, Paul A Lyons, Michael P Weekes

## Abstract

Certain serum proteins, including CRP and D-dimer, have prognostic value in patients with SARS-CoV-2 infection. Nonetheless, these factors are non-specific, and provide limited mechanistic insight into the peripheral blood mononuclear cell (PBMC) populations which drive the pathogenesis of severe COVID-19. To identify novel cellular phenotypes associated with disease progression, we here describe a comprehensive, unbiased analysis of the total and plasma membrane proteomes of PBMCs from a cohort of 40 unvaccinated individuals with SARS-CoV-2 infection, spanning the whole spectrum of disease severity. Combined with RNA-seq and flow cytometry data from the same donors, we define a comprehensive multi-omic profile for each severity level, revealing cumulative immune cell dysregulation in progressive disease. In particular, the cell surface proteins CEACAMs1, 6 and 8, CD177, CD63 and CD89 are strongly associated with severe COVID-19, corresponding to the emergence of atypical CD3^+^CD4^+^CD177^+^ and CD16^+^CEACAM1/6/8^+^ mononuclear cells. Utilisation of these markers may facilitate real-time patient assessment by flow cytometry, and identify immune cell populations that could be targeted to ameliorate immunopathology.

## Introduction

SARS-CoV-2 continues to present a public health crisis due to a slow global rollout of vaccination programmes and emergence of novel virus variants. Viral pathogenesis comprises an initial stage of virus replication followed by immune cell recruitment and cytokine production. Most infected individuals generate an effective immune response that achieves viral clearance without excessive tissue damage, presenting with no or mild symptoms. However, a minority experience severe disease, driven by a dysregulated and hyperactive immune response and characterised by high levels of pro-inflammatory mediators such as IL-6 and TNFα^1,2^. The consequent enhanced vascular permeability, thrombosis and tissue damage can lead to severe pneumonia, acute respiratory distress syndrome, multiple organ failure and death^3,4^.

Several studies have aimed to define perturbations in circulating immune cell subsets and secreted factors during severe COVID-19, utilising techniques such as bulk and single cell transcriptomics, cytometry panels and plasma proteomics. These have shown that severe COVID-19 is associated with profound peripheral lymphopenia^5–8^ charactersised by recruitment of NK cells to the lung tissue from peripheral blood, and expansion of activated yet functionally impaired^9,10^ inflammatory NK cells expressing cytotoxic factors and interferon-stimulated genes^10–12^. Unusually for a viral infection, profound peripheral neutrophilia is also observed^13,14^ due to release of immature, inflammatory neutrophils via emergency myelopoiesis and a hyperinflammatory phenotype reminiscent of bacterial sepsis^15–17^. Circulating CD4^+^ and CD8^+^ T-cells are depleted^5,7,8,18^, concurrent with expansion of activated effector cell subpopulations^9,19^, and express high levels of exhaustion markers such as PD-1 and TIM3^9,19,20^. Significant changes are observed in the myeloid compartment, such as depletion of non-classical CD16^+^ monocytes^17,21,22^ and expansion of CD14^+^HLA-DR^low^ cells resembling immunosuppressive monocytes observed in sepsis^17^. In addition, platelets exhibit a hyperactivated phenotype defined by expression of the activation marker CD62P (P-selectin) and readily form pro-thrombotic platelet-leukocyte aggregates^23–25^.

A comprehensive understanding of the immune dysregulation and immunopathology that underpins COVID-19 is essential to identify patients at risk of progressing to severe disease in order to provide early therapeutic intervention. Here, we performed a detailed, unbiased proteomic analysis of the peripheral blood mononuclear cell (PBMC) plasma membrane and cellular proteomes from seven healthy controls and 33 unvaccinated individuals with acute SARS-CoV-2 infection across the spectrum of COVID-19 disease. These data complement previous characterisation of the same cohort by whole blood transcriptomics and cytometric phenotyping^5^ alongside PBMC single-cell sequencing^26^. We identified progressive upregulation of a novel group of proteins expressed by multiple immune cell subpopulations, including CD4^+^ T-cells and non-classical monocytes, as disease severity increased. These data further our understanding of immune dysfunction in COVID-19 and define a novel set of cellular phenotypes that could be used to identify patients at risk of progression to severe disease.

## Results

### Patient cohort and workflow

SARS-CoV-2 PCR-positive subjects were recruited to this study as part of a larger cohort by Bergamaschi et al between 31st March and 20th July 2020. Subjects with no or mild symptoms were recruited from routine screening of healthcare workers (HCWs). Patients with COVID-19 were recruited at or soon after admission to Cambridge University Hospitals or the Royal Papworth Hospital. Participants were divided into five categories of clinical severity (**Figures 1A-B, Table S1A**): (A) asymptomatic HCWs; (B) HCWs who were either still working with mild symptoms insufficient to meet criteria for self-isolation or were symptomatic and self-isolating;^27,28^ (C) patients who presented to hospital but never required oxygen supplementation; (D) patients who were admitted to hospital and whose maximal respiratory support was supplemental oxygen; and (E) patients who at some point required assisted ventilation. One patient who died without admission to intensive care were also included in this severe group. Additionally, seven healthy controls (HCs), distributed across a range of age and gender, were included.

**Figure 1.**
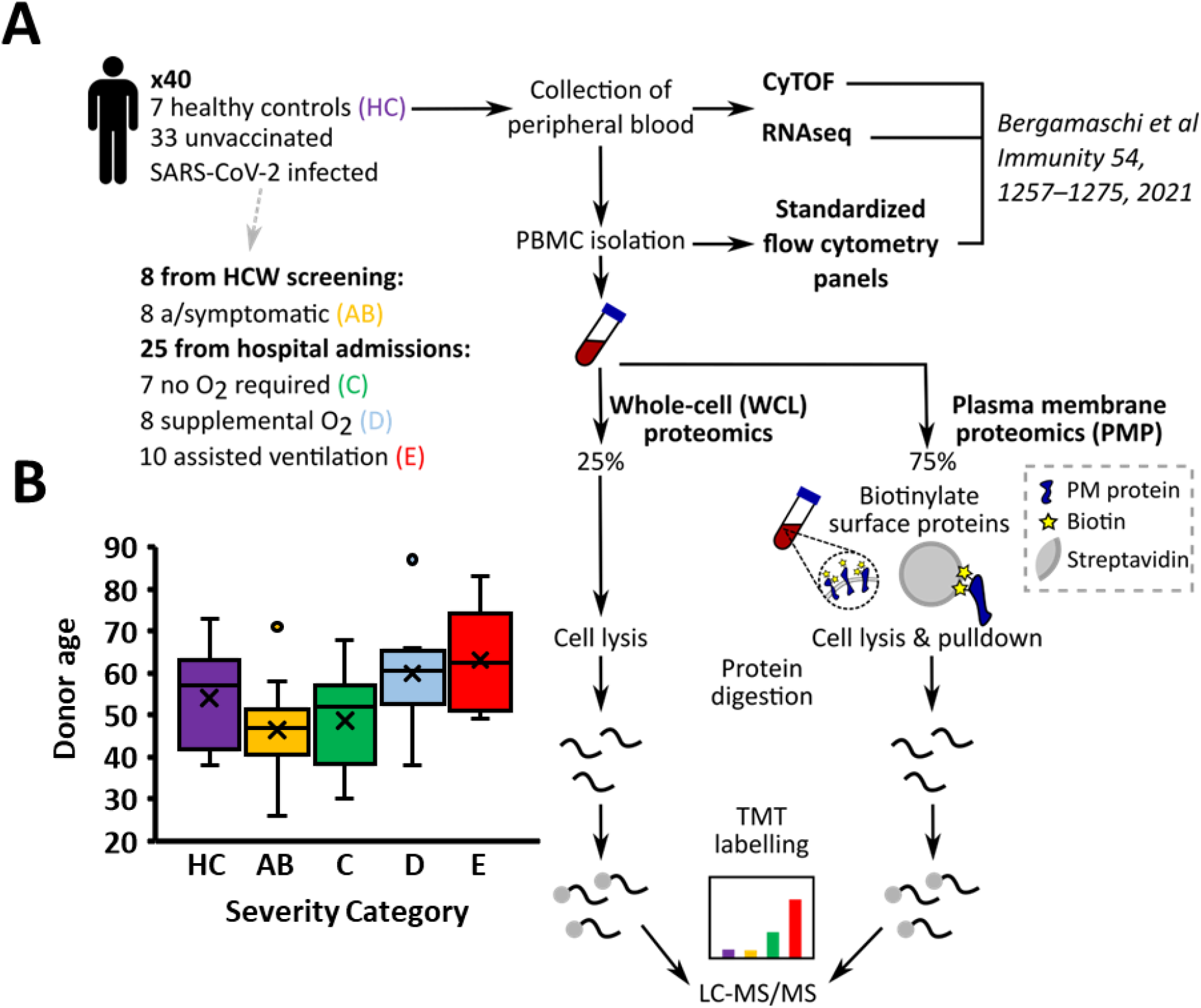
Cohort outline and experimental workflow. (A) Details of study participants (B) Distribution of participant age across disease severity categories (C) Schematic of experimental workflow. Patient PBMC samples were thawed and processed in parallel for TMT-based multiplexed analysis of whole-cell and plasma-membrane proteomes.

Peripheral blood mononuclear cells (PBMCs) were characterised using two parallel workflows for proteomic analysis, to profile changes in the whole-cell (WCL) and plasma membrane (PM) proteomes. Peptide samples were labelled with TMT reagents and analysed in a series of multiplexed tandem mass spectrometry (MS) experiments (**Figure 1A**)^29^. Aliquots of the same samples were previously analysed using flow cytometry panels and RNAseq^5^.

### Identification of cellular phenotypes associated with disease severity by mass-spectrometry

7704 proteins were quantified in the WCL dataset and 597 proteins were quantified in the PM dataset. Statistical analyses indicated that there were no significant differences in protein abundance between donors with asymptomatic or mild symptomatic disease (classes A & B) and these data were therefore combined for further analysis. In whole cell lysates, differential protein expression was most pronounced in severe COVID-19 (classes D & E). Whereas 45 and 82 proteins were upregulated >2-fold relative to healthy controls in class AB and C donors, strikingly, 278 and 392 proteins were upregulated >2-fold in classes D and E, respectively. These included several proteins previously identified as upregulated in severe COVID-19: C-reactive protein (CRP), complement component 9 (C9) and lipopolysaccharide binding protein (LBP) (**Figures 2A-B**). Gene Ontology (GO) analysis identified significant enrichment of terms associated with antiviral defence in class AB donors, including upregulation of multiple interferon stimulated genes such as IFITs, Mx proteins and OAS proteins. By comparison, proteins highly upregulated in classes D & E were enriched in functions related to antimicrobial and antibacterial defence (**Figure 2B, Table S2**). Data from all proteomic experiments in this study are shown in **Table S3**. Here, the worksheet ‘‘Lookup’’ is interactive, enabling generation of graphs of expression of any of the proteins quantified.

**Figure 2.**
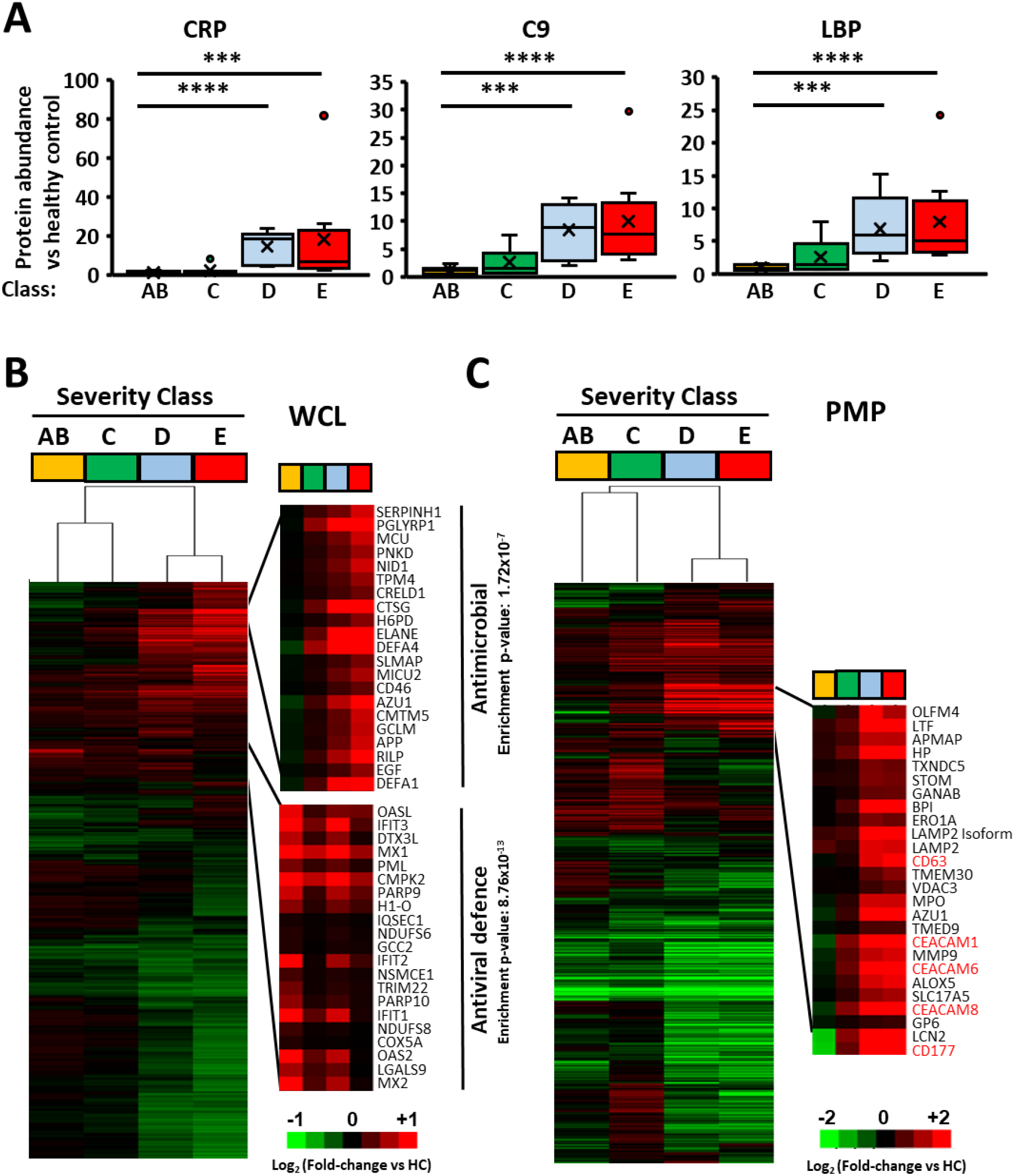
Severe COVID is associated with substantial changes in both the whole-cell and plasma membrane PBMC proteomes. (A) Examples of data for three proteins quantified in our analysis that have been reported to be upregulated in PBMC from individuals with severe COVID-19. Ordinary one-way ANOVA with Tukey’s multiple comparisons post-hoc test on Log_2_-transformed data: *p<0.05, **p<0.005, ***p<0.0005, ****p<0.0001. (B) Hierarchical cluster analysis of fold change in expression of 5,226 whole cell proteins quantified across all three WCL analyses for each severity class compared to healthy control. Fold changes were derived from data averaged across all donors from a given disease severity class in comparison to average data from healthy controls analysed in the same mass spectrometry experiment. Enlargements of two subclusters are shown, highlighting groups of proteins that were upregulated in classes D and E vs AB (top panel) or in classes AB vs E (bottom panel). DAVID enrichment terms and corresponding Benjamini-Hochberg-corrected p-values are shown for each cluster. (C) Hierarchical cluster analysis of 522 plasma-membrane-enriched proteins quantified in both PM analyses. Enlargement of one subcluster is shown, identifying a group of proteins that are highly upregulated at the PBMC cell-surface in severe COVID-19. Proteins that best discriminate between mild and severe disease on the basis of principal component analysis (**Figure S1**) are highlighted in red.

To identify cell surface phenotypes associated with COVID-19 severity, which could also be readily utilised in diagnostic or therapeutic settings, equivalent analyses were conducted on PM data (**Figure 2C**). Overall, 54 and 49 proteins were upregulated >2-fold in classes D and E versus healthy control, including a cluster of proteins that were specifically upregulated in severe, but not mild, disease (**Figure 2C**, right panel). Interestingly, this group of proteins included several members of the carcinoembryonic antigen-related cell adhesion molecule (CEACAM) family, CEACAMs 1, 6 and 8, in addition to immunological markers CD177, CD63 and CD89. Principal component analysis (PCA) of WCL and PM data confirmed that samples from classes D & E clustered separately from healthy controls and classes A, B & C (**Figures S1A-B**). Analysis of PCA loadings revealed that a substantial proportion of the variation between classes resulted from changes in CEACAM proteins, CD177, CD63 and CD89 (**Figures S1C-D**). Further analysis of protein profiles for each of these candidate markers identified a consistent and significant upregulation in marker abundance with increasing disease severity in both WCL and PM data (**Figure 3**). This trend was also observed for corresponding genes in parallel samples analysed by whole-blood RNA-seq by Bergamaschi et al^5^ (**Table S1B**), further supporting the proteomic conclusions (**Figure 3**). As a result, this group of markers were selected for further validation and characterisation.

**Figure 3.**
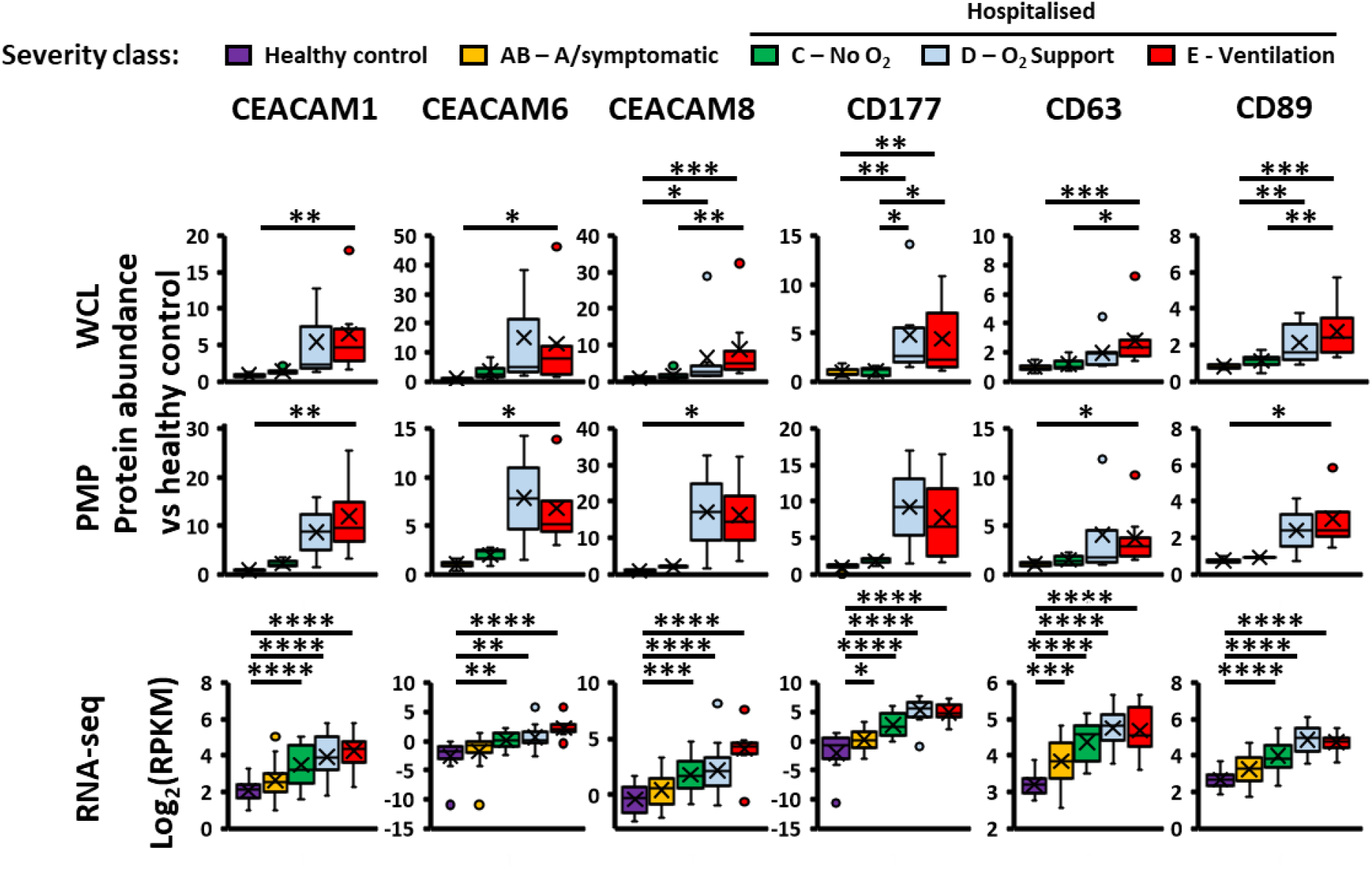
Identification of host proteins associated with severe COVID using complementary datasets. Protein and RNA expression data for candidate markers identified in our analysis as being upregulated in PMBC from individuals with severe COVID-19. RNA expression data at the earliest available timepoint was extracted from previous whole-blood RNAseq analysis of a larger cohort including our donors^5^ and expressed in Log2(RPKM). Ordinary one-way ANOVA with Tukey’s multiple comparisons post-hoc test on Log_2_-transformed data: *p<0.05, **p<0.005, ***p<0.0005, ****p<0.0001.

**Figure S1.**
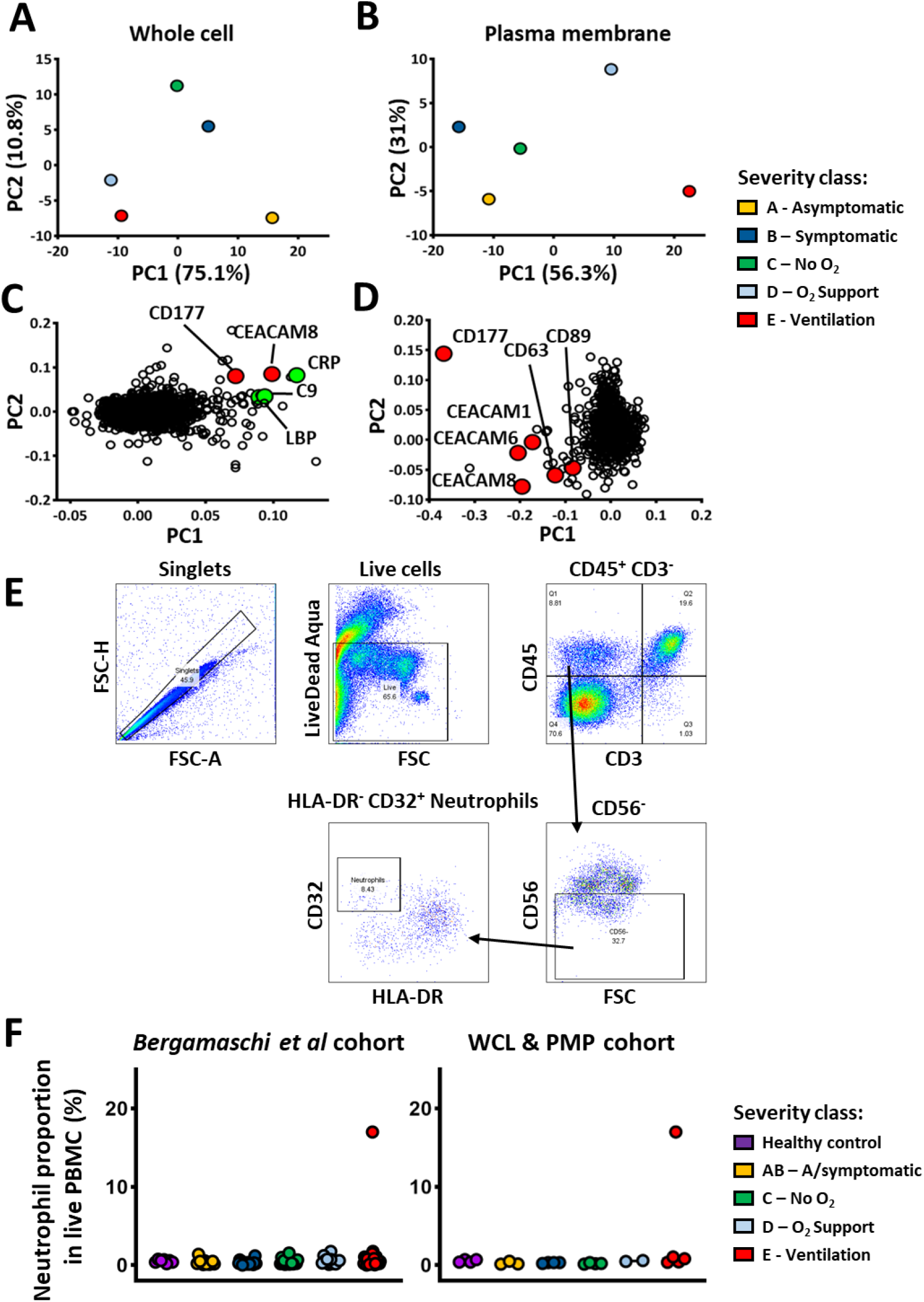
Principal component analysis (PCA) of proteomic data, and assessment of PBMC sample neutrophil contamination. (A) PCA of 5,226 whole-cell protein abundance ratios relative to healthy control for each disease severity class, derived from proteins quantified across all three WCL analyses. (B) PCA of 522 plasma membrane protein abundance ratios relative to healthy control for each disease severity class, derived from proteins quantified across both PM analyses. (C) Individual protein loadings for the WCL PCA in Fig. S1A. Proteins previously identified as upregulated in severe disease are highlighted in green, with novel candidate markers highlighted in red. (D) Individual protein loadings for the PM PCA in Fig. S1B. Novel candidate markers are highlighted in red. (E) Gating strategy to define the neutrophil population in a re-analysis of flow cytometry data from Bergamaschi et al^5^. (F) Flow cytometry data showing the proportion of neutrophils in live PBMC as a percentage for donations from Bergamaschi et al^5^ (left panel) or from the subset of these donations selected for proteomic analysis (right panel).

Of note, CEACAM8 and CD177 are classically regarded as neutrophil markers. As neutrophilia is a hallmark of severe COVID-19, we first determined whether observations resulted from neutrophil contamination of PBMC samples, despite exclusion of granulocytes during PBMC density gradient isolation. PBMC immunophenotyping flow cytometry data were reanalysed for all donors in the larger cohort from which proteomic samples were drawn (**Table S1C**)^5^. Mature neutrophil contamination was only present in a single sample, indicating an absence of confounding systemic contamination (**Figures S1E-F**). Notably, the one donor with identified mature neutrophil sample contamination (CV0144, class E) did not exhibit greater upregulation of CEACAM8 or CD177 versus other class E donors, suggesting that upregulation of these markers is neutrophil-independent.

### Validation and phenotyping of identified markers by flow-cytometry

To verify candidate markers and to phenotype cell populations expressing these proteins, multicolour flow cytometry was performed on a cohort of 36 donors across the range of disease severity classes (**Figures 4A, S2A-B, Table S1D**). Corresponding to previous observations, this revealed a progressive and significant decrease in lymphocytes with worsening COVID-19, concurrent with a significant increase in platelet abundance and no overall trend in the myeloid compartment (**Figure S2C**). Detailed phenotyping identified further changes associated with disease severity in specific subpopulations, including depletion of CD56^+^ NK cells, CD3^+^CD4^+^ and CD8^+^ T-cells and increases in both resting CD62P^-^ and activated CD62P^+^ platelets (**Figure 4B**), again consistent with previously documented changes in circulating immune cells during acute COVID-19^6–8,17^. A trend towards depletion of CD3^+^γδTCR^+^ T-cells and increased abundance of CD16^+^ non-classical monocytes was also observed, although due to sample limitations it was not possible to collect sufficient events required for statistical significance.

**Figure 4.**
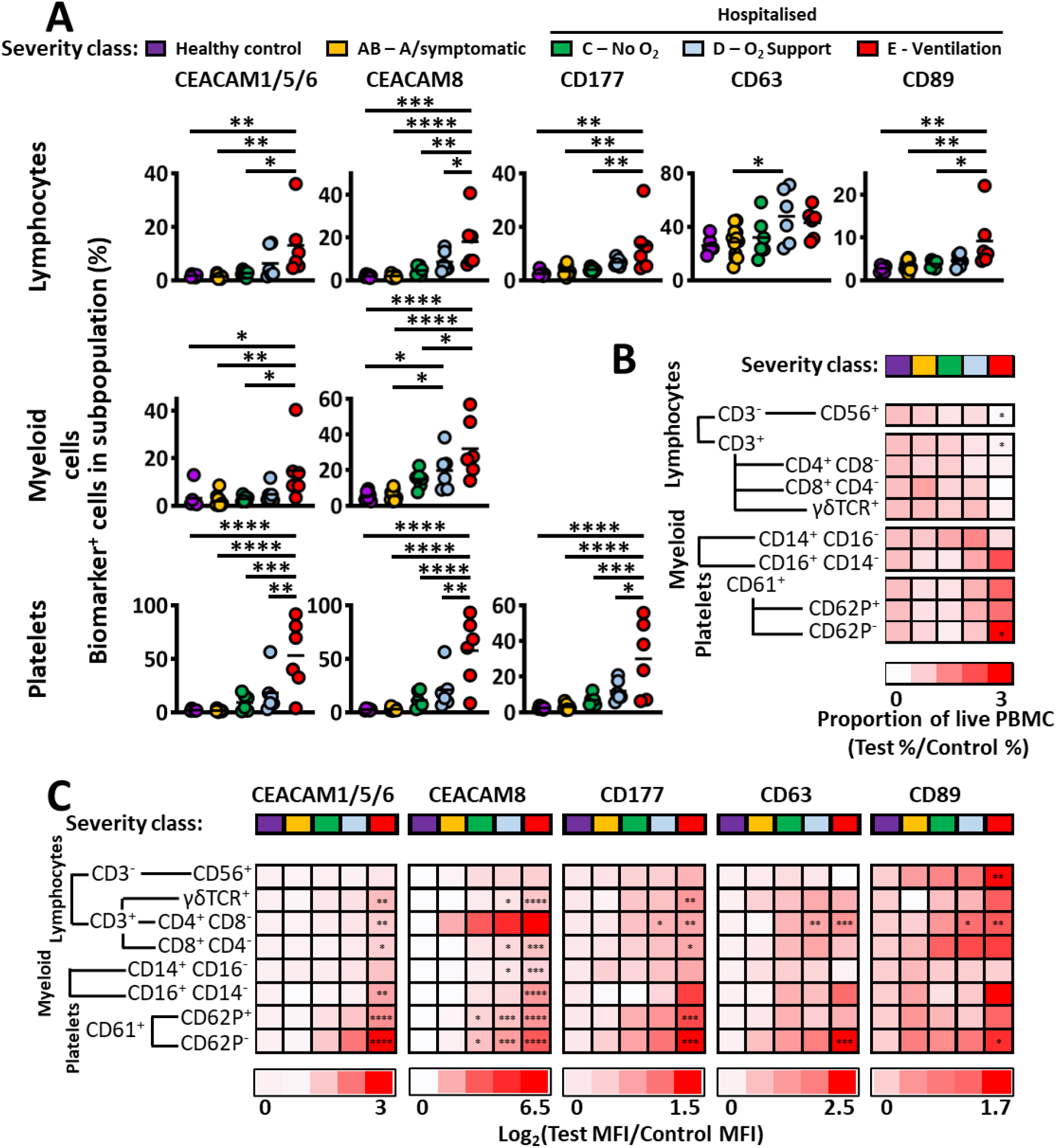
Validation and characterisation of candidate markers of disease severity by flow cytometry. (A) Flow cytometry data for candidate markers, showing the proportion of target-positive cells within a live PBMC subpopulation (gating strategy – **Figure S2A**). Ordinary one-way ANOVA with Tukey’s multiple comparisons post-hoc test. *p<0.05, **p<0.005, ***p<0.0005, ****p<0.0001. (B) Heatmap of PBMC composition across disease severity classes, expressed as ratio comparing the proportion of each cell population in live PBMC relative to healthy controls (gating strategy – **Figure S3B**). p-values: as Figure 4A. (C) Heatmap of biomarker expression across immune cell subsets, expressed as log_2_ (fold change in geometric mean fluorescence intensity relative to healthy controls). p-values: as Figure 4A.

**Figure S2.**
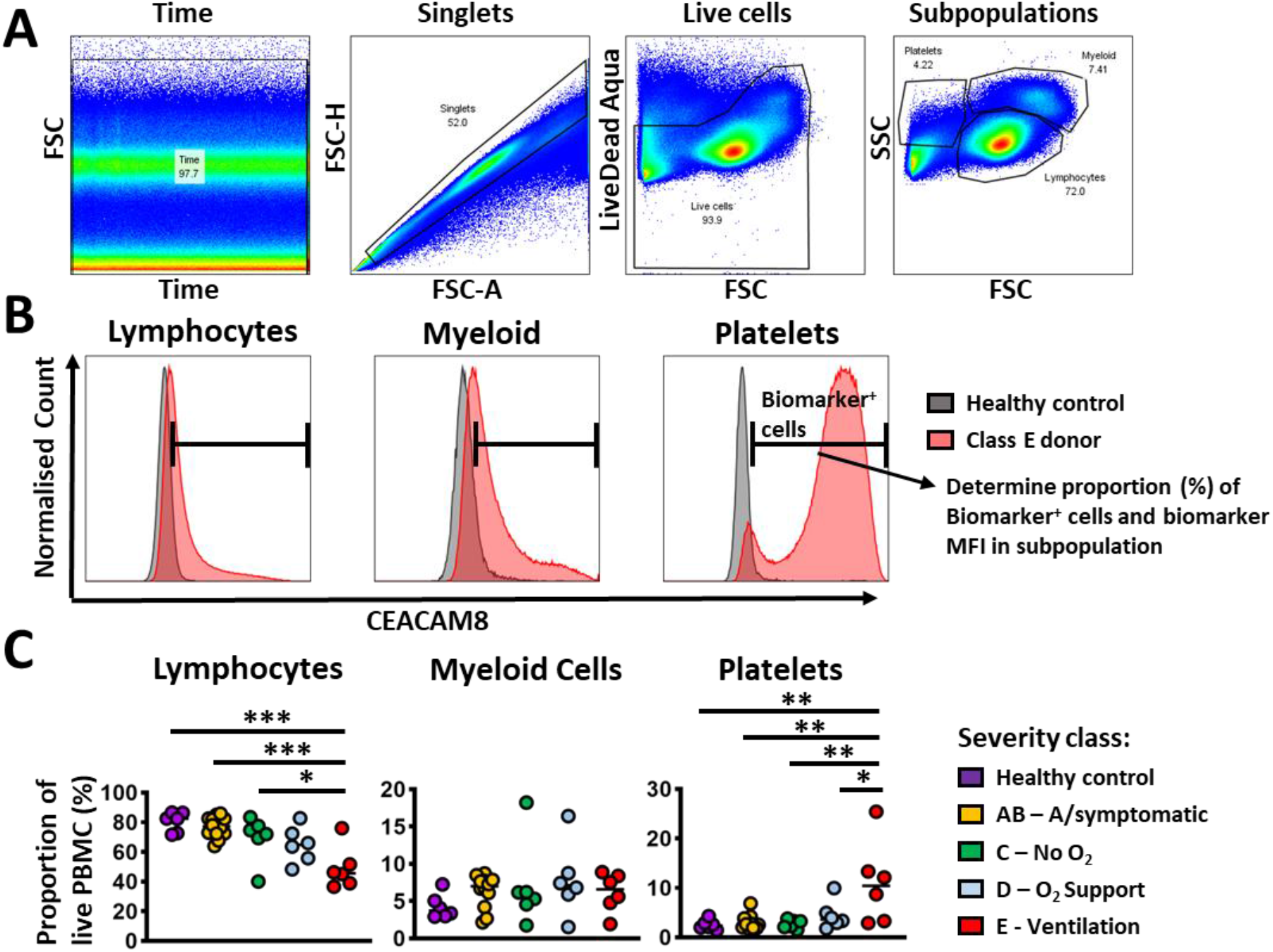
Flow cytometry gating methodology and proportions of PBMC subpopulations. (A) The flow cytometry gating strategy for selection of live singlets and definition of PBMC subpopulations. (B) Methodology for determining the proportion of marker-positive cells and measuring marker geometric mean fluorescence intensity (MFI) in each subpopulation. (C) The proportions of lymphocytes, myeloid cells and platelets in PBMC from each severity class. Ordinary one-way ANOVA with Tukey’s multiple comparisons post-hoc test. *p<0.05, **p<0.005, ***p<0.0005, ****p<0.0001.

Interestingly, candidate markers exhibited expression changes associated with disease severity in bulk lymphocyte, myeloid and platelet populations. The frequency and intensity of CEACAM1, 6 and 8 expression was significantly upregulated on lymphocyte and myeloid cell populations in class E donors compared to both healthy controls and patients with mild disease (**Figures 4A, S3A**) and on platelets, significantly upregulated in severe disease versus healthy controls and patients with mild or moderate disease. Notably, frequency and intensity of CEACAM8 expression on myeloid cells was also significantly upregulated on patients with moderate versus mild disease, indicating progressive upregulation of this marker with increasing disease severity. The frequency and intensity of CD177 expression was upregulated in severe disease on both lymphocytes and platelets, CD89 on lymphocytes alone and the intensity of CD63 and CD89 expression was significantly increased on platelets (**Figures 4A, S3A**).

Further phenotyping was performed to define cell populations upregulating candidate markers (**Figure S3B**). Among lymphocytes, CD3^+^CD4^+^ T-cells significantly upregulated CEACAM1/6, CD177, CD63 and CD89 (**Figure 4C**), in addition to strongly upregulating CEACAM8, although substantial variation between donors was observed. CD3^+^CD8^+^ T-cells and γδT-cells significantly upregulated CD177 and CEACAMs 1, 6 and 8, while CD56+ NK cells upregulated CD89 alone. Within the myeloid compartment, significant upregulation of CEACAM8 was observed on CD14^+^ classical monocytes and upregulation of CEACAMs 1, 6 and 8 on non-classical CD16^+^ monocytes, although low cell counts confounded interpretation of data for this population. Resting CD61^+^CD62P^-^ platelets significantly upregulated all identified markers in patients with severe disease and activated CD61^+^CD62P^+^ platelets upregulated CEACAMs 1, 6 and 8 in addition to CD177. Combined tSNE analysis of live PBMC sampled from all class E donors (**Figure S4A)** indicated that each identified biomarker was co-upregulated on distinct, singular CD4^+^ T-cell (**Figure S4B**), CD62P^-^ platelet (**Figure S4C**) and CD16^+^ monocyte populations (**Figure S4D**).

**Figure S3.**
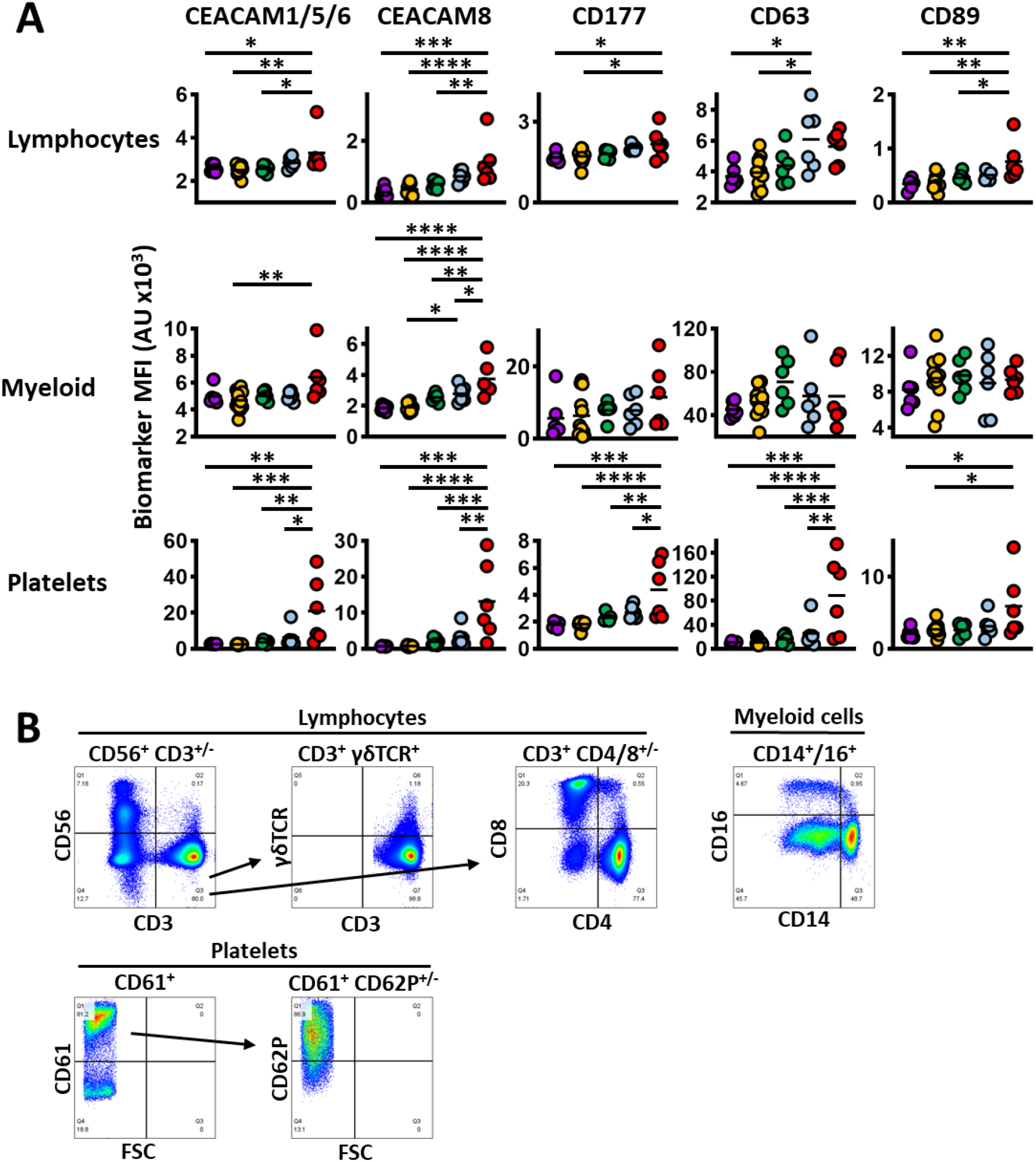
Validation and characterisation of candidate phenotypes associated with disease severity by flow cytometry. (A) Flow cytometry data, showing geometric mean fluorescence intensity in arbitrary units of fluorescence (AU). Ordinary one-way ANOVA with Tukey’s multiple comparisons post-hoc test. *p<0.05, **p<0.005, ***p<0.0005, ****p<0.0001. (B) Gating strategy for definition of immune cell subsets within PBMC subpopulations.

**Figure S4.**
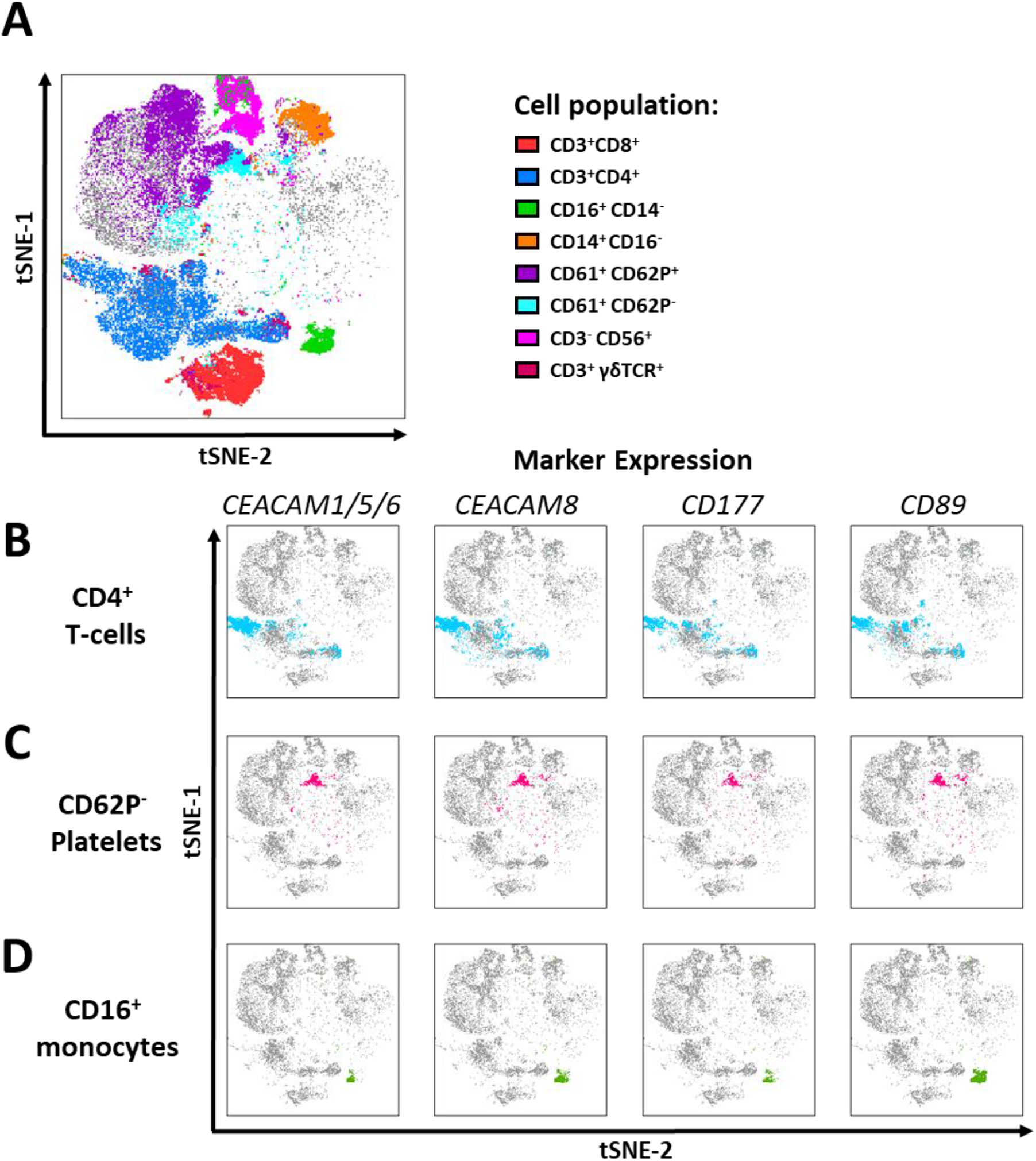
Co-upregulation of markers on CD4^+^ T-cells, CD62P^-^ platelets and CD16^+^ cells. (A) tSNE visualisation of 6×10^5^ live PBMC sampled from each class E donor, coloured by cell population (B) Overlay of marker-positive CD4^+^ T-cells onto tSNE analysis in S4A (C) Overlay of marker-positive CD62P^-^ platelets onto tSNE analysis in S4A (D) Overlay of marker-positive CD16^+^ cells onto tSNE analysis in S4A

## Discussion

Understanding the complex immunobiology of COVID-19 is essential in developing predictive measures both of the severity of acute disease and to predict the development and progress of long COVID. This knowledge will also be vital to understanding efficacy of novel therapies. Here we present a searchable analysis of the PBMC cellular and plasma membrane proteomes during acute SARS-CoV-2 infection in a cohort of donors spanning the spectrum of COVID-19 disease. Our data indicate a profound shift in PBMC proteome profiles from mild to severe disease, echoing observations made in the whole blood transcriptome^5^ and plasma proteome^30^ and reflecting the significant remodelling of circulating immune cell composition during COVID-19. Notably, we observed highly significant enrichment of terms related to microbial defence among cellular proteins upregulated during severe disease. Patient metadata indicated that only a small number of patients in classes D and E had confirmed secondary infections, further corroborating the emergence of a sepsis-like state driving COVID-19 immunopathology^17^. In addition, selective upregulation of canonical interferon-stimulated genes such as the IFIT and Mx families was observed in patients with mild disease. Of note, time-normalised transcriptomic analysis of the wider cohort from which our samples derived^5^ suggested that PBMC from patients with severe disease that are collected relatively early after symptom onset can also exhibit upregulated ISG expression which subsequently wanes. We observed a similar trend for donors sampled in the earlier phases of infection (Tables S1, S3).

Unbiased profiling of the PBMC plasma membrane proteome identified a unique signature of severe COVID-19, marked by the upregulated expression of a group of proteins with immunoregulatory functions: CEACAM1, 6 and 8, CD177, CD63 and CD89. CD177 is a glycosyl-phosphatidylinositol (GPI)-linked surface glycoprotein that is canonically regarded as a neutrophil marker^31^, although expression on monocytes has also been reported^32^. CD177 plays characterised roles in mediating neutrophil endothelial transmigration via binding to PECAM1^33^ in addition to activation and degranulation as part of a CD177/PR3-CD11b/CD18 complex^34,35^. CD63 is a ubiquitously expressed member of the tetraspanin membrane protein family involved in cell adhesion and intracellular trafficking^36^. Typically localised to intracellular compartments, cell surface CD63 is used as a marker of platelet^37^ and T-cell^38^ activation in addition to granulocyte degranulation^39,40^. CD89 is an Fc receptor expressed on neutrophils and monocytes^32^ that binds to IgA immune complexes^41^ and CRP^42^, initiating cell activation, cytokine release^43^ and has a role in protecting against bacterial sepsis^44^.

CEACAMs 1, 6 and 8 belong to a family of immunoglobulin-like surface glycoproteins that can form homophilic and heterophilic interactions in conjunction with an array of binding partners that participate in a diverse range of processes, including cell adhesion, signalling and immunoregulation^45^. Members of the CEACAM family are expressed on a broad range of cell types: CEACAM1 on epithelial cells, endothelial cells^45^ and activated T-cells^46^; CEACAM6 on epithelial cells, neutrophils and monocytes^32^; CEACAM8 on granulocytes and monocytes^32^. Notably, members of the family have established roles in modulating immune cell function. CEACAM1 has an established inhibitory role in T-cells^47^, suppressing signalling through the TCR-CD3 complex via phosphatase recruitment^48^ in response to stimuli such as homophilic interaction^49^ with CEACAM1 on antigen-presenting cells or heterophilic interaction with CEACAM6^50^. In addition, CEACAM1 forms a complex with and mediates the inhibitory function of the exhaustion marker TIM3^51^ on T-cells and negatively regulates NK cell activity^52^. CEACAM8 is upregulated on granulocytes following activation^53^, mediates adhesion to endothelial cells^54^, cytokine release^55^ and is highly upregulated on neutrophils in bacterial sepsis^56^.

Interestingly, CD177 levels have previously been associated with COVID-19 severity and ICU admission in an analysis of a French cohort by whole-blood transcriptomics and serum profiling^57^. The authors proposed that increased circulating CD177 reflects the dysregulated neutrophil activation observed in severe COVID-19. We also observed an equivalent progressive and significant upregulation of CD177 in PBMC preparations. Isolation of PBMCs by density gradient centrifugation typically excludes granulocytes and an absence of mature neutrophil contamination was verified by flow cytometry. Phenotyping indicated the progressive emergence of a CD3^+^CD4^+^CD177^+^ T-cell population as disease severity increased in the context of a broader depletion of both CD4^+^ and CD8^+^ T-cells. This population also co-upregulated CEACAMs 1 and 6, CD63, CD89 and likely CEACAM8, although the latter marker exhibited substantial inter-donor variation.

Simultaneous expression of CEACAMs 6 and 8, CD177 and CD89 on CD4^+^ T-cells is intriguing, as expression of these markers is regarded as restricted to granulocytes and monocytes and represents a unique phenotype that has not been previously reported. CD177 expression may plausibly facilitate migration into critical tissues and cell activation in the context of infection. Upregulation of CD89 is particularly interesting due to its role in bacterial sepsis^44^ and the relationship between severe COVID-19 and a sepsis-like syndrome. Recognition of IgA^41^, the most prevalent antibody isotype in the respiratory tract, and CRP^42^, a widely used biomarker of severe COVID-19, by CD89 may have functional implications in cytokine release from this cell population. Concurrent upregulation of the activation factors CEACAM8 and CD89 and the inhibitory factors CEACAM1 and 6 is unusual and resembles observations of SARS-CoV-2-specific CD8^+^ T-cells in patients with severe disease expressing both exhaustion markers and transcriptional signatures of inflammatory cytokine production and cytotoxicity^58,59^. Our data suggest that a non-canonical but similar phenotype may therefore also develop in the CD4^+^ T-cell population in the context of severe COVID-19.

It is presently unclear whether this CD4^+^ population contains SARS-CoV-2-specific cells, represents activated bystander T-cells or cells with other specificities. SARS-CoV-2-specific CD4^+^ T-cells have been observed from 2-4 days after symptom onset^60^, consistent with the timescale in our cohort. Antigen-independent activation of CD8^+^ T-cell populations by inflammatory cytokines has been reported in several viral infections, including hepatitis A^61^, influenza A^62^ and SARS-CoV-2^5^ and are proposed to contribute to immunopathology in these settings. Bystander activation of CD4^+^ T-cells is also proposed to contribute to pathogenesis during herpes simplex virus^63,64^ and dengue virus infection^65^. Regardless of antigen specificity, this CD4^+^ T-cell population may plausibly constitute a further example of immune dysregulation in severe COVID-19 and contribute to immunopathology.

Previous studies have identified a hyperactive platelet phenotype in critically ill patients with COVID-19 that is proposed to contribute to hypercoagulability and tissue damage observed during acute respiratory distress syndrome. This phenotype is characterised by upregulated expression of the activation markers CD62P, LAMP3 and CD63^24,66^, increased formation of platelet-leukocyte aggregates^24,25^ and indications of degranulation^67^. Supporting these observations, we observed a clear shift in platelet phenotype in patients with severe COVID-19, with a profound increase in both activated CD61^+^CD62P^+^ and non-activated CD61^+^CD62P^-^ platelets and upregulated expression of CD63. Interestingly, we also identified a CD62P^-^ platelet population that co-upregulated CD177, CD89 and CEACAMs 1, 6 and 8 to high levels. Expression of CD89, for example, may facilitate platelet degranulation. Upregulation of CD177 and CEACAM family members is particularly interesting in this context, given their roles in cell adhesion through homophilic or heterophilic interactions. Expression of these factors on platelets, monocytes and neutrophils may represent a mechanism that facilitates cell-cell contacts and formation of thrombotic aggregates.

As in other studies^17,21^, our data indicated perturbations in the myeloid compartment. Phenotyping indicated that CEACAMs 1, 6 and 8 were prominently co-upregulated on a small population of CD16^+^CD14^-^ cells and CEACAM8 on CD14^+^ CD16^-^ cells. One possible explanation for the former observation is that the identified proteins are upregulated on non-classical CD16^+^ monocytes. Several studies have described depletion of non-classical monocytes in severe COVID-19^17,21,22^, although this was not observed in single-cell sequencing of our cohort^26^ and does not preclude expansion of a subpopulation within a broader population contraction. Alternatively, our data may represent increased abundance of immature low-density pro- or pre-neutrophils (LDNs) during severe disease. LDNs migrate with PBMC during density gradient isolation unlike their mature counterparts and are typically released via emergency myelopoiesis^68^ during infection^69,70^, sepsis^71,72^ and in autoimmune conditions^73^. LDNs are associated with a dysfunctional immunosuppressive environment^74,75^. Emergence of immature neutrophils has previously been reported in severe COVID-19^15,17,21^, including a CEACAM8^+^ subpopulation^17^, although this population exhibited low to intermediate CD16 expression^15^. Prior characterisation of PBMC from this cohort by single-cell RNAseq documented the appearance of a rare C1QA/B/C^+^CD16^+^ monocyte population in patients with severe disease that is predicted to interact with and contribute to platelet activation^26^, but did not observe emergence of any mature or immature neutrophil population. Our data may therefore suggest the appearance of a CD16^+^CEACAM1/6/8^+^ monocyte subset in the context of severe COVID-19.^26^. Taken together, our observations of expression of CEACAM family members and CD177 on platelets and CEACAM expression on both classical and non-classical monocytes may represent another mechanism for formation of pathological platelet-monocyte aggregates.

In summary, we have identified novel immune-cell surface phenotypes associated with COVID-19 severity, and have characterised unusual CD4 T-cell, resting platelet and monocyte populations that co-express individual marker proteins in PBMC from individuals with severe COVID-19. These cellular phenotypes could be readily utilised in identifying individuals with, or developing severe disease in addition to providing future avenues for expanding understanding of mechanisms of immunopathology. Further longitudinal studies are required to determine the prognostic value of these markers and their potential involvement with persistent ‘long COVID’ symptoms.

## STAR Methods

### Key resources table

| REAGENT or RESOURCE | SOURCE | IDENTIFIER |
| --- | --- | --- |
| Antibodies |  |  |
| Anti-human CD61 BUV395 (RUU-PL7F12) | BD | AB_2871633 |
| Anti-human CD16 BUV496 (3G8) | BD | AB_2870224 |
| Anti-human CD4 BUV563 (RPA-T4) | BD | AB_2870854 |
| Anti-human CD62P BUV737 (AK-4) | BD | AB_2873476 |
| Anti-human CD8 BUV805 (RPA-T8) | BD | AB_2873737 |
| Anti-human CD66a/c/e BV605 (ASL-32) | BioLegend | AB_2800910 |
| Anti-human CD3 BV785 (OKT3) | BioLegend | AB_2563507 |
| Anti-human CD177 FITC (MEM-166) | BioLegend | AB_2072603 |
| Anti-human CD66b PE (G10F5) | BioLegend | AB_2077857 |
| Anti-human CD56 PE-Dazzle (HCD56) | BioLegend | AB_2563564 |
| Anti-human CD63 APC (H5C6) | BioLegend | AB_10916521 |
| Anti-human CD89 AlexaFluor 700 (A59) | BioLegend | AB_2750067 |
| Anti-human CD14 APC-Fire (M5E2) | BioLegend | AB_2632660 |
| Chemicals, peptides, and recombinant proteins |  |  |
| TMTpro 16plex Label Reagent Set | Thermo Fisher Scientific | Cat# A44522 |
| HPLC water | VWR | Cat# 23595.328 |
| LC-MS grade acetonitrile | Merck | Cat# 1.00029.2500 |
| Formic acid | Thermo Fisher Scientific | Cat# 85178 |
| Guanidine hydrochloride (8M) | Thermo Fisher Scientific | Cat# 24115 |
| DL-Dithiothreitol | Sigma-Aldrich | Cat# 43815-1G |
| Iodoacetamide | Sigma | Cat# I1149-5G |
| Lysyl Endopeptidase | Wako | Cat# 125-02543 |
| Trypsin Protease | Pierce | Cat# 90058 |
| Hydroxylamine | Sigma | Cat# 438227 |
| Acetonitrile, Extra Dry | Acros Organics | Cat# AC364311000 |
| HEPES (1M, pH 7.0-7.6) | Sigma | Cat# H0887 |
| LIVE/DEAD Fixable Aqua Dead Cell Stain Kit | Thermo Fisher Scientific | Cat# L34966 |
| Human TruStain FcX | Biolegend | Cat# 422302 |
| BD Horizon Brilliant Stain Buffer | BD | Cat# 563794 |
| FluoroFix Buffer | BioLegend | Cat# 422101 |
| Critical commercial assays |  |  |
| Micro BCA Protein Assay Kit | Thermo Fisher Scientific | Cat# 23235 |
| AbC Total Antibody Compensation Bead Kit | Thermo Fisher Scientific | Cat# A10497 |
| ArC Amine Reactive Compensation Bead Kit | Thermo Fisher Scientific | Cat# A10346 |
| Deposited data |  |  |

| Software and algorithms |  |  |
| --- | --- | --- |
| "MassPike", a Sequest-based software pipeline for quantitative proteomics | Professor Steven Gygi's lab, Harvard Medical School, Boston, USA | N/A |
| Cluster 3.0 | Stanford University, University of Tokyo | http://bonsai.hgc.jp/%7Emdehoon/software/cluster/software.htm |
| Java Treeview | SourceForge.net | http://jtreeview.sourceforge.net/ |
| Perseus | Max Planck Institute of Biochemistry, Martinsried, Germany | https://maxquant.net/perseus/ |
| DAVID software | Huang da et al., 2009 | https://david.ncifcrf.gov/ |
| Graphpad Prism 9 | GraphPad Software | https://www.graphpad.com/scientific-software/prism/ |
| R | R Core Team 2015 | N/A |
| RStudio Desktop 2022.02.2+485 | RStudio | https://www.rstudio.com/ |
| FlowJo | BD | v10,<br>https://www.flowjo.com/solutions/flowjo |
| Other |  |  |
| Orbitrap Fusion Lumos Mass Spectrometer | Thermo Fisher Scientific | Cat# IQLAAEGAAP FADBMBHQ |
| Dionex UltiMate 3000 UHPLC | Thermo Fisher | N/A |
| Cytek Aurora Flow Cytometer | Cytek Biosciences | N/A |

### Resource Availability

#### Lead Contact

Further information and requests for resources and reagents should be directed to and will be fulfilled by the lead contact, Prof. Michael Weekes.

#### Materials availability

This study did not generate new unique reagents.

#### Data and code availability

The mass spectrometry raw files and associated unmodified peptide and protein quantitation data have been deposited in the ProteomeXchange Consortium via the iProX partner repository with identifier IPX0005417000^76,77^.

### Experimental Model and Subject Details

#### Human subjects

25 patients with confirmed or suspected SARS-CoV-2 infection were recruited between 31/3/2020 and 20/7/2020 from Cambridge University Hospitals (CUH) or the Royal Papworth Hospital (Cambridge, UK)^5,78,79^. Infection was confirmed in 24 participants via a nucleic acid amplification tests. One donor presented with relevant symptoms and required low flow oxygen support but tested negative by PCR. 15 individuals were recruited through the Cambridge University Hospitals healthcare worker (HCW) PCR screening programme^27,28^, including eight with no or mild symptoms and a positive test for SARS-CoV-2, and seven who tested negative for SARS-CoV-2 (Figure 1). These negative control samples were selected to represent the age and sex distribution of SARS-CoV-2-positive participants. Recruitment of inpatients and HCWs at Cambridge University Hospitals was undertaken by the NIHR Cambridge Clinical Research Facility outreach team and the NIHR BioResource research team. Ethical approval was obtained from the East of England – Cambridge Central Research Ethics Committee (‘‘NIHR BioResource’’ REC ref 17/EE/0025, and ‘‘Genetic variation AND Altered Leucocyte Function in health and disease - GANDALF’’ REC ref 08/H0308/176). One healthy control donor was obtained for flow cytometry analysis under ethical approval from the Health Research Authority (HRA) Cambridge Central Research Ethics Committee (97/092). All participants provided informed consent. Detailed information on the patients can be found in **Table S1**.

### Method Details

#### Clinical data collection

Clinical data were collected from medical charts and entered into spreadsheets. Available laboratory test results were extracted from Epic electronic health records (Cambridge University Hospitals) and from MetaVision ICU (Royal Papworth hospital ITU). SARS-CoV-2-positive HCW participants were categorised into two groups according to whether they were asymptomatic (group A) or had COVID-19 symptoms at the time of PCR testing (group B). Symptoms considered to be possible manifestations of COVID-19 were new onset fever (>37.8 °C), cough, loss of sense of small, hoarseness, nasal discharge or congestion, shortness of breath, wheeze, headache, muscle aches, nausea, vomiting and diarrhoea^28^.

Hospital patients were assigned to one of three groups, reflecting the maximum level of respiratory support received during their hospital stay. Group C patients did not receive any supplemental oxygen. Group D patients received supplemental oxygen using low flow nasal prongs, a simple face mask, a Venturi mask or a non re-breathe face mask. Patients who received any of non-invasive ventilation (NIV), mechanical ventilation or extracorporeal membrane oxygenation (ECMO) were assigned to group E. Patients in group D who died in hospital during the study were also assigned to group E. In patients who were already established on home NIV for chronic respiratory failure, NIV delivered as per the home prescription (e.g. nocturnal) was not considered for the purpose of classification. Moreover, oxygen requirements that were clearly not related to COVID-19 were also not considered for classification purposes.

#### Peripheral blood mononuclear cell preparation and flow immunophenotyping

Each participant provided 27 mL of peripheral venous blood collected into 9 mL sodium citrate tubes. Peripheral blood mononuclear cells (PBMCs) were isolated using Leucosep tubes (Greiner Bio-One) with Histopaque 1077 (Sigma) by centrifugation at 800 × g for 15 minutes at room temperature. PBMCs at the interface were collected, rinsed twice with autoMACS running buffer (Miltenyi Biotech) and cryopreserved in FBS with 10% DMSO. All samples were processed within 4 hours of venepuncture.

For proteomic and flow cytometry analysis, frozen PBMC samples were thawed in a water bath at 37°C and immediately diluted in TexMACS media (Miltenyi Biotech), centrifuged, resuspended in TexMACS supplemented with 10U/ml DNAse (Benzonase, Merck-Millipore) and rested at 37°C for 1h. PBMCs were then centrifuged, resuspended in fresh media and counted. For proteomic analysis, PMBCs were washed twice in ice-cold PBS pH 7.4 (Sigma). 75% of cells were used for plasma membrane profiling, while the remaining 25% was used for whole cell lysate proteomic analysis. For flow cytometry, cells were directly processed as described below.

#### Plasma membrane profiling

Following washing, cellular surface sialic acid residues were oxidised then biotinylated with 1 mM sodium meta-periodate (Thermo), 100mM aminooxy-biotin (Biotium) and 10 mM aniline (Sigma) in ice cold PBS pH 6.7, by rocking the cells with 3 mL of the mix at 4 °C for 30 minutes. The reaction was quenched by adding glycerol (Sigma) to a final concentration of 1 mM. The cells were then washed twice with ice-cold PBS pH 7.4 containing CaCl_2_ and MgCl_2_, and then lysed with 1.6% Triton X-100 (Thermo), 150 mM NaCl (Sigma), 1 × protease inhibitor (complete, without EDTA (Roche)), 5 mM iodoacetamide (Sigma) and 10 mM Tris-HCl pH 7.6 (Sigma) for 30 minutes at 4 °C. Nuclei and debris were removed by centrifugation at 4 °C, once at 4,000 × g for 5 minutes then twice at 13,000 × g for 5 minutes. Samples were then snap-frozen in liquid nitrogen and stored at – 80 °C prior to immunoprecipitation and protein digestion.

The following washes were performed using Poly-Prep columns (Bio-Rad) attached to a vacuum manifold. Biotinylated proteins were enriched by incubating with high affinity streptavidin agarose beads (Pierce) at 4 °C for 75 minutes on a rotor, then were washed extensively with lysis buffer (1% Triton X-100, 150 mM NaCl, 10 mM Tris-HCl pH 7.6) and PBS (with CaCl_2_ and MgCl_2_) / 0.5% SDS (Invitrogen). Beads were next incubated with PBS / 0.5% SDS / 100 mM dithiothreitol (DTT) for 20 minutes at room temperature. Further washes were performed with UC buffer (6 M urea in 0.1 M Tris-HCl pH 7.6), before alkylation with UC buffer containing 50 mM iodoacetamide for 20 minutes at room temperature in the dark. Beads were washed again with UC buffer and HPLC-grade H_2_O, transferred to a screw cap column (Pierce) and then proteins were digested on-bead with 35 µl of 8 ng/µl trypsin (Thermo) in 200 mM HEPES pH 8.5 (Sigma) for 3 h in a shaking 37 °C incubator. The digested peptides were eluted and stored at -80 °C before TMT labelling.

#### Whole cell lysate digestion

After washing cells were lysed in 50 µl of 6 M guanidine (Thermo) / 50 mM HEPES pH 8.5, vortexed extensively and sonicated. Cell debris was removed by centrifuging twice at 13,000 × g for 10 minutes at 4 °C.

DTT was added to a final concentration of 5 mM and incubated at room temperature for 20 mins. Cysteine residues were alkylated with 15 mM iodoacetamide and incubated for 20 min at room temperature in the dark. Excess iodoacetamide was quenched with DTT for 15 mins. Samples were diluted with 200 mM HEPES pH 8.5 to a final concentration of 1.5 M guanidine, followed by digestion at room temperature for 3 h with LysC protease (Wako) at a 1:100 protease-to-protein ratio. Samples were further diluted with 200 mM HEPES pH 8.5 to a concentration of 0.5 M guanidine. Trypsin was then added at a 1:100 protease-to-protein ratio followed by overnight incubation at 37°C. The reaction was quenched with 5% formic acid and centrifuged at 21,000 × g for 10 min to remove undigested protein. Peptides were subjected to C18 solid-phase extraction (SPE, Sep-Pak, Waters) and vacuum-centrifuged to near-dryness.

#### Peptide labelling with Tandem Mass Tags

TMTpro reagents (0.8 mg, Thermo) were dissolved in 45 µl anhydrous acetonitrile. For PM samples, 35 µl of digested peptide was labelled with 10 µl of TMT reagent in a final volume of 50 µl with a final acetonitrile concentration of 30% (v/v). For WCL samples, desalted peptides were dissolved in 200 mM HEPES pH 8.5 and peptide concentration measured by microBCA (Thermo). 30 µg of peptide was labelled with 10 µl of TMT reagent at a final acetonitrile concentration of 30% (v/v).

PM and WCL samples were then processed in parallel. Following incubation with TMT reagents at room temperature for 1 h, the reactions were quenched with hydroxylamine to a final concentration of 0.5% (v/v). Equivalent amounts of each sample were combined in batches. An unfractionated 1h analysis of each batch was carried out initially to ensure equivalent peptide loading across each TMT channel, thus avoiding the need for excessive electronic normalisation. Samples were then subjected to high pH reversed-phase (HpRP) fractionation (below).

#### Offline HpRp fractionation

TMT-labelled tryptic peptides derived from WCL samples were fractionated using an Ultimate 3000 RSLC UHPLC system (Thermo) equipped with a 2.1 mm internal diameter (ID) x 25 cm long, 1.7 µm particle Kinetix Evo C18 column (Phenomenex). Mobile phase consisted of A: 3% acetonitrile (MeCN), B: MeCN, and C: 200 mM ammonium formate pH 10. Isocratic conditions were 90% A/10% C, and C was maintained at 10% throughout the gradient elution. Separations were conducted at 45 °C. Samples were loaded at 200 µl/minute for 5 min. The flow rate was then increased to 400 µl/minute over 5 minutes, after which the elution gradient proceeded as follows: 0-19% B over 10 minutes, 19-34% B over 14.25 minutes, 34-50% B over 8.75 minutes, followed by a 10 min wash at 90% B. UV absorbance was monitored at 280 nm and 15 s fractions were collected into 96 well microplates using the integrated fraction collector. For WCL samples, fractions were recombined orthogonally in a checkerboard fashion, combining alternate wells from each column of the plate into a single fraction, and commencing combination of adjacent fractions in alternating rows. This yielded two sets of 12 combined fractions, A and B. For PM samples, all wells in sets of two adjacent columns were recombined to yield six combined fractions. Wells were excluded prior to the start or after the cessation of elution of peptide-rich fractions, as identified from the UV trace. Samples were dried in a vacuum centrifuge and resuspended in 10 µl MS solvent (4% MeCN/5% formic acid) prior to LC-MS3.

For WCL samples, a series of 12 or 24 LC-MS3 analyses were next performed (either all of set A or all of sets A and B). For PM samples, either six fractions and an unfractionated singleshot sample were analysed (batch 1) or an unfractionated singleshot sample was analysed (batch 2).

#### LC-MS3

Mass spectrometry data were acquired using an Orbitrap Lumos (Thermo Fisher Scientific, San Jose, CA). An Ultimate 3000 RSLC nano UHPLC equipped with a 300 µm ID x 5 mm Acclaim PepMap µ-Precolumn (Thermo) and a 75 µm ID x 50 cm 2.1 µm particle Acclaim PepMap RSLC analytical column was used. Loading solvent was 0.1% formic acid (FA), analytical solvent A: 0.1% FA and B: 80% MeCN + 0.1% FA. All separations were carried out at 55°C. Samples were loaded at 5 µl/min for 5 min in loading solvent before beginning the analytical gradient. The following gradient was used: 3-7% B over 3 min, 7-37% B over 173 min, followed by a 4 min wash at 95% B and equilibration at 3% B for 15 min. Each analysis used a MultiNotch MS3-based TMT method (4, 5). The following settings were used: MS1: 380-1500 Th, 120,000 Resolution, 2×10^5^ automatic gain control (AGC) target, 50 ms maximum injection time. MS2: Quadrupole isolation at an isolation width of m/z 0.7, CID fragmentation (normalised collision energy (NCE) 35) with ion trap scanning in turbo mode from m/z 120, 1.5×10^4^ AGC target, 120 ms maximum injection time. MS3: In Synchronous Precursor Selection mode the top 6 MS2 ions were selected for HCD fragmentation (NCE 65) and scanned in the Orbitrap at 60,000 resolution with an AGC target of 1×10^5^ and a maximum accumulation time of 150 ms. Ions were not accumulated for all parallelisable time. The entire MS/MS/MS cycle had a target time of 3 s. Dynamic exclusion was set to ± 10 ppm for 70 s. MS2 fragmentation was trigged on precursors 5×10^3^ counts and above.

#### Flow cytometry

1×10^6^ cells per donor were resuspended in 100µl Horizon Brilliant Stain Buffer (BD) and blocked with Human TruStain FcX (BioLegend) at room temperature for 5 minutes. Cells were then stained with a 15-colour panel (see Key Resources Table) for 30 minutes at 4°C, before washing in an excess of PBS, centrifugation and resuspension in 100µl FluoroFix Buffer (BioLegend). Single colour compensation controls were prepared for the panel using AbC Compensation beads (Thermo) and ArC Compensation beads (Thermo) for antibody stains and amine-reactive viability stains respectively, or healthy control PBMCs. Samples were then analysed on a Cytek Aurora flow cytometer (Cytek Biosciences) and data analysed in FlowJo (BD).

### Quantification and Statistical Analysis

#### Data Analysis

Mass spectra were processed using a Sequest-based software pipeline for quantitative proteomics, ‘‘MassPike’’, through a collaborative arrangement with Professor Steven Gygi’s laboratory at Harvard Medical School. MS spectra were converted to mzXML using an extractor built upon Thermo Fisher’s RAW File Reader library (version 4.0.26). In this extractor, the standard mzxml format has been augmented with additional custom fields that are specific to ion trap and Orbitrap mass spectrometry and essential for TMT quantitation. These additional fields include ion injection times for each scan, Fourier Transform-derived baseline and noise values calculated for every Orbitrap scan, isolation widths for each scan type, scan event numbers and elapsed scan times. This software is a component of the MassPike software platform and is licensed by Harvard Medical School.

A combined database was constructed from the human Uniprot database (11/11/2020), the SARS-CoV-2 Uniprot reference proteome and common contaminants such as porcine trypsin and endoproteinase LysC. The combined database was concatenated with a reverse database composed of all protein sequences in reversed order. Searches were performed using a 20 ppm precursor ion tolerance. Fragment ion tolerance was set to 1.0 Th. TMT tags on lysine residues and peptide N termini and carbamidomethylation of cysteine residues (57.02146 Da) were set as static modifications, while oxidation of methionine residues (15.99492 Da) was set as a variable modification.

To control the fraction of erroneous protein identifications, a target-decoy strategy was employed ^80^. Peptide spectral matches (PSMs) were filtered to an initial peptide-level false discovery rate (FDR) of 1% with subsequent filtering to attain a final protein-level FDR of 1%. PSM filtering was performed using a linear discriminant analysis, as described previously ^80^. This distinguishes correct from incorrect peptide IDs in a manner analogous to the widely used Percolator algorithm (https://noble.gs.washington.edu/proj/percolator/), though employing a distinct machine-learning algorithm. The following parameters were considered: XCorr, DCn, missed cleavages, peptide length, charge state, and precursor mass accuracy. Protein assembly was guided by principles of parsimony to produce the smallest set of proteins necessary to account for all observed peptides (algorithm described in ^80^).

Proteins were quantified by summing TMT reporter ion counts across all matching peptide-spectral matches using “MassPike”, as described previously^81^. Briefly, a 0.003 Th window around the theoretical m/z of each reporter ion was scanned for ions and the maximum intensity nearest to the theoretical m/z was used. The primary determinant of quantitation quality is the number of TMT reporter ions detected in each MS3 spectrum, which is directly proportional to the signal-to-noise (S:N) ratio observed for each ion. Conservatively, every individual peptide used for quantitation was required to contribute sufficient TMT reporter ions so that each on its own could be expected to provide a representative picture of relative protein abundance ^81^. A per-sample S:N ratio of >15 was required such that, for example, for a 16-plex experiment a combined S:N ratio of >240 across all TMT reporter ions would be needed for a peptide to pass filtering. An isolation specificity filter with a cut-off of 50% was additionally employed to minimise peptide co-isolation ^81^. Peptides meeting the stated criteria for reliable quantitation were then summed by parent protein, in effect weighting the contributions of individual peptides to the total protein signal based on their individual TMT reporter ion yields. Protein quantitation values were exported for further analysis in Excel.

For protein quantitation, reverse and contaminant proteins were removed, then each reporter ion channel was summed across all quantified proteins and normalised assuming equal protein loading across all channels. Missing values in the mass spectrometry data were imputed for a small number of donor-protein datapoints (11 datapoints in total for all WCL analyses, two datapoints in total for both PM analyses) by setting missing values to the minimum intensity observed for the protein within each multiplexed experiment. Data for all HLA isoforms were removed, due to variation in HLA allele expression between donors. Proteins originating from red blood cell (RBC) contaminants were removed if they met two criteria: (1) they were identified as one of the top 15% most abundant RBC proteins from Ravenhill et al^82^ and (2) they had a coefficient of variation greater than 0.75 in any given disease severity class when comparing protein abundance fold-change versus healthy control across donors.

Hierarchical centroid clustering of proteomic data was carried out in Cluster 3.0 (Stanford University) using an uncentred Pearson correlation similarity metric.

#### Statistical Analysis

For displayed proteins or cell populations, ordinary one-way ANOVA tests with Tukey’s adjustment for multiple comparisons were carried out in GraphPad Prism 9 on log_2_-transformed fold-change values for each donor versus healthy controls (Figs. 2A, 3A, 4C) or on untransformed cell population proportion values (Fig. 4A, 4B, S3A). Adjusted p-values <0.05 were considered significant. Table S1: one- and three-way ANOVAs with Tukey’s multiple comparisons post-hoc test for WCL MS data or Kruskal-Wallis tests for PM MS data were carried out in Perseus.

#### Pathway Analysis

The Database for Annotation, Visualization and Integrated Discovery (DAVID) was used to identify enrichment of pathways in upregulated gene clusters as specified in the text. In each case, the cluster was searched against the background of all proteins quantified in our proteomics data using default settings.

#### tSNE Analysis

Live PBMC (gated as in S2A) from class E donors CV0191, CV0197, CV0203, CV0212, CV0272 and CV0278 were concatenated, downsampled and tSNE analysis carried out in FlowJo v10. The tSNE parameters utilised were as follows: learning configuration: opt-SNE, iterations: 1000, perplexity: 30, learning rate: 438000, KNN algorithm: Exact (vantage point tree) and gradient algorithm: Barnes-Hut.

## Data Availability

The mass spectrometry raw files and associated unmodified peptide and protein quantitation data have been deposited in the ProteomeXchange Consortium via the iProX partner repository with identifier IPX0005417000.

## Acknowledgments

We are grateful to Prof. Steve Gygi for providing access to the “MassPike” software pipeline for quantitative proteomics. We thank NIHR BioResource volunteers for their participation, and gratefully acknowledge NIHR BioResource centres, NHS Trusts and staff for their contribution. We thank the National Institute for Health and Care Research, NHS Blood and Transplant, and Health Data Research UK as part of the Digital Innovation Hub Programme. This research was funded by a grant from the Addenbrooke’s Charitable Trust and a Medical Research Council (MRC) Project Grant (MR/W025647/1) to MPW, a Wellcome Investigator Award (200871/Z/16/Z) and an MRC Programme Grant (MR/L019027) to KGCS, an MRC studentship (MR/N013433/1) to AFE, grants from the MRC (MR/S036113/1), CRUK (C1163/A21762) and the Aging Biology Foundation to FCN and BG, grants from the MRC (MR/S000081X/1) and Wellcome Trust (WT/204870/Z/16/Z) to MRW, an MRC Transition Support Fellowship (MR/T032413/1) and NHSBT grant (WPA15-02) to NJM, and the Wellcome Trust Institutional Strategic Support Fund (204845/Z/16/Z) to MPW and NJM. This study was additionally supported by the Cambridge Biomedical Research Centre, UK, CVC Capital Partners and the Evelyn Trust (20/75). This research was facilitated by the CIMR Flow Cytometry Core Facility, the NIHR Cambridge Biomedical Research Centre Cell Phenotyping Hub, the NIHR BioResource, the NIHR Cambridge Biomedical Research Centre and the NIHR Cambridge Clinical Research Facility. The views expressed are those of the author(s) and not necessarily those of the NIHR or the Department of Health and Social Care. For the purpose of open access, the author has applied a CC BY public copyright licence to any Author Accepted Manuscript version arising from this submission.

## The CITIID-NIHR BioResource COVID-19 Collaboration

Stephen Baker^2,6^, John Bradley^1,3,6,11,15^, Patrick Chinnery^3,23,24^, Daniel Cooper^11, 25^, Gordon Dougan^2,6^, Ian Goodfellow^7^, Ravindra Gupta^2,6,13,16^, Nathalie Kingston^3,4^, Paul J. Lehner^2,6,13^, Paul A. Lyons^2,6^, Nicholas J. Matheson^2,6,13,33^, Caroline Saunders^9^, Kenneth G. C. Smith^2,6^, Charlotte Summers^6,12,26^, James Thaventhiran^19^, M. Estee Torok ^6,13,14^, Mark R. Toshner^6,8,26^, Michael P. Weekes^2,6,13,34^, Christoph Hess^2,6^, Gisele Alvio^9^, Sharon Baker^9^, Areti Bermperi^9^, Karen Brookes^9^, Ashlea Bucke^9^, Jo Calder^9^, Laura Canna^9^, Cherry Crucusio^11^, Isabel Cruz^9^, Ranalie de Jesus^9^, Katie Dempsey^9^, Giovanni Di Stephano^9^, Jason Domingo^9^, Anne Elmer^9^, Julie Harris^29^, Sarah Hewitt^9^, Heather Jones^9^, Sherly Jose^9^, Jane Kennet^9^, Yvonne King^29^, Jenny Kourampa^9^, Emily Li^6^, Caroline McMahon^9^, Anne Meadows^9^, Vivien Mendoza^9^, Criona O’Brien^29^, Charmain Ocaya^9^, Ciro Pasquale^9^, Marlyn Perales^9^, Jane Price^9^, Rebecca Rastall^9^, Carla Ribeiro^9^, Jane Rowlands^9^, Valentina Ruffolo^9^, Hugo Tordesillas^9^, Phoebe Vargas^9^, Bensi Vergese^9^, Laura Watson^9^, Jieniean Worsley^9^, Julie-Ann Zerrudo^9^, Laura Bergamashi^2,6^, Ariana Betancourt^2,6^, Georgie Bower^2,6^, Ben Bullman^2^, Chiara Cossetti^34^, Aloka De Sa^6^, Benjamin J. Dunmore^6^, Maddie Epping^2,6^, Stuart Fawke^34^, Stefan Gräf ^3,6^, Richard Grenfell^36^, Andrew Hinch^37^, Josh Hodgson^6^, Christopher Huang^6^, Oisin Huhn^38^, Kelvin Hunter^2,6^, Isobel Jarvis^6^, Emma Jones^39^, Maša Josipović^32^, Ekaterina Legchenko^6^, Daniel Lewis^6^, Joe Marsden^40^, Jennifer Martin^6^, Federica Mescia^2,6^, Ciara O’Donnell^6^, Ommar Omarjee^6^, Marianne Perera^6^, Linda Pointon^6^, Nicole Pond^2,6^, Nathan Richoz^6^, Nika Romashova^6,37^, Natalia Savoinykh^37^, Rahul Sharma^6^, Joy Shih^6^, Mateusz Strezlecki^36^, Rachel Sutcliffe^6^, Tobias Tilly^6^, Zhen Tong^6^, Carmen Treacy^6^, Lori Turner^2,6^, Jennifer Wood^6^, Marta Wylot^41^, John Allison^3,4^, Heather Biggs^3,18^, John R. Bradley^1,3,6,11,15^, Helen Butcher^3,5^, Daniela Caputo^3,5^, Matt Chandler^3,5^, Patrick Chinnery^3,23,24^, Debbie Clapham-Riley^3,5^, Eleanor Dewhurst^3,5^, Christian Fernandez^3,^ Anita Furlong^3,5^, Barbara Graves^3,5^, Jennifer Gray^3,5^, Sabine Hein^3,5^, Tasmin Ivers^3,5^, Emma Le Gresley^3,5^, Rachel Linger^3,5^, Mary Kasanicki^3,11^, Rebecca King^3,5^, Nathalie Kingston^3,4^, Sarah Meloy^3,5^, Alexei Moulton^3,5^, Francesca Muldoon^3,5^, Nigel Ovington^3,4^, Sofia Papadia^3,5^, Christopher J. Penkett^3,4^, Isabel Phelan^3,5^, Venkatesh Ranganath^3,4^, Roxana Paraschiv^3,4^, Abigail Sage^3,5^, Jennifer Sambrook^3,4^, Ingrid Scholtes^3,5^, Katherine Schon^3,17,18^, Hannah Stark^3,5^, Kathleen E. Stirrups^3,4^, Paul Townsend^3,4^, Neil Walker^3,4^, Jennifer Webster^3,5^, Mayurun Selvan^35^, Petra Polgarova^12^,Sarah L. Caddy^2,6^, Laura G. Caller^20,21^, Yasmin Chaudhry^7^, Martin D. Curran^22^, Theresa Feltwell^6^, Stewart Fuller^20^, Iliana Georgana^7^, Grant Hall^7^, William L. Hamilton^6,13,14^, Myra Hosmillo^7^, Charlotte J. Houldcroft^6^, Rhys Izuagbe^7^, Aminu S. Jahun^7^, Fahad A. Khokhar^2,6^, Anna G. Kovalenko^7^, Luke W. Meredith^7^, Surendra Parmar^22^, Malte L. Pinckert^7^, Anna Yakovleva^7^, Emily C. Horner^19^, Lucy Booth^19^, Alexander Ferreira^19^, Rebecca Boston^19^, Robert Hughes^19^, Juan Carlos Yam Puc^19^, Nonantzin Beristain-Covarrubias^19^, Maria Rust^19^, Thevinya Gurugama^19^, Lihinya Gurugama^19^, Thomas Mulroney^19^, Sarah Spencer^19^, Zhaleh Hosseini^19^, Kate Williamson^19^.

^1^NIHR Cambridge Biomedical Research Centre, Cambridge Biomedical Campus, Cambridge, UK

^2^Cambridge Institute of Therapeutic Immunology and Infectious Disease (CITIID), Jeffrey Cheah Biomedical Centre, Cambridge Biomedical Campus, Cambridge, UK

^3^NIHR BioResource, Cambridge University Hospitals NHS Foundation Trust, Cambridge Biomedical Campus, Cambridge, UK

^4^Department of Haematology, School of Clinical Medicine, University of Cambridge, Cambridge Biomedical Campus, Cambridge, UK

^5^Department of Public Health and Primary Care, School of Clinical Medicine, University of Cambridge, Cambridge Biomedical Campus, Cambridge, UK

^6^Department of Medicine, School of Clinical Medicine, University of Cambridge, Cambridge Biomedical Campus, Cambridge, UK

^7^Division of Virology, Department of Pathology, University of Cambridge, Cambridge, UK

^8^Royal Papworth Hospital NHS Foundation Trust, Cambridge, UK

^9^Cambridge Clinical Research Centre, Addenbrooke’s Hospital, Cambridge University Hospitals NHS Foundation Trust, Cambridge, UK

^10^Intensive Care Unit, Royal Papworth Hospital NHS Foundation Trust, Cambridge, UK

^11^Addenbrooke’s Hospital, Cambridge University Hospitals NHS Foundation Trust, Cambridge Biomedical Campus, Cambridge, UK

^12^Intensive Care Unit, Addenbrooke’s Hospital, Cambridge University Hospitals NHS Foundation Trust, Cambridge Biomedical Campus, Cambridge, UK

^13^Department of Infectious Diseases, Addenbrooke’s Hospital, Cambridge University NHS Hospitals Foundation Trust, Cambridge, UK

^14^Department of Microbiology, Addenbrooke’s Hospital, Cambridge University NHS Hospitals Foundation Trust, Cambridge, UK

^15^Department of Renal Medicine, Addenbrooke’s Hospital, Cambridge University Hospitals NHS Foundation Trust, Cambridge, UK

^16^Africa Health Research Institute, Durban, South Africa

^17^Clinical Genetics, Addenbrooke’s Hospital, Cambridge University Hospitals NHS Foundation Trust, Cambridge, UK

^18^Department of Clinical Neurosciences, School of Clinical Medicine, University of Cambridge, Cambridge Biomedical Campus, Cambridge, UK

^19^MRC Toxicology Unit, Gleeson Building, Tennis Court Road, Cambridge, UK

^20^University of Cambridge, Cambridge, UK

^21^The Francis Crick Institute, London, UK

^22^Public Health England, Clinical Microbiology and Public Health Laboratory, Cambridge, UK

^23^Department of Clinical Neurosciences, School of Clinical Medicine, University of Cambridge, Cambridge Biomedical Campus, Cambridge, UK

^24^Medical Research Council Mitochondrial Biology Unit, Cambridge Biomedical Campus, Cambridge, UK

^25^Global and Tropical Health Division, Menzies School of Heath Research and Charles Darwin University, Darwin, Northern Territory, Australia

^26^Heart and Lung Research Institute, Cambridge Biomedical Campus, Cambridge, UK

^27^Department of Rheumatology, Addenbrooke’s Hospital, Cambridge University Hospitals NHS Foundation Trust, Cambridge, UK

^28^Cambridge Cancer Trials Centre, Addenbrooke’s Hospital, Cambridge University Hospitals NHS Foundation Trust, Cambridge, UK

^29^Department of Paediatrics, University of Cambridge, Cambridge Biomedical Campus, Cambridge, UK

^30^Patient Safety, Addenbrooke’s Hospital, Cambridge University Hospitals NHS Foundation Trust, Cambridge, UK

^31^Clinical Research Network: Eastern, Addenbrooke’s Hospital, Cambridge University Hospitals NHS Foundation Trust, Cambridge, UK

^32^Institute of Metabolic Science, Addenbrooke’s Hospital, Cambridge University Hospitals NHS Foundation Trust, Cambridge, UK

^33^NHS Blood and Transplant, Cambridge, UK

^34^Cambridge Institute for Medical Research, Biomedical Campus, Hills Rd, Cambridge UK

^35^ Department of Respiratory Medicine, Cambridge University Hospitals NHS Foundation Trust, Cambridge, UK

^36^Cancer Research UK Cambridge Institute, University of Cambridge, Cambridge, UK

^37^NIHR Cambridge Biomedical Research Centre Cell Phenotyping Hub, Cambridge Biomedical Campus. Cambridge, UK

^38^Department of Obstetrics & Gynaecology, The Rosie Maternity Hospital, Robinson Way, Cambridge, UK

^39^Department of Veterinary Medicine, Cambridge, UK

^40^Department of Chemical Engineering and Biotechnology, University of Cambridge, Cambridge, UK

^41^Department of Biochemistry, University of Cambridge, Cambridge, UK

### Supplemental tables

**Table S1**

(A) Details of donors used for proteomic analysis

(B) Details of donors used to generate whole-blood RNA-seq data in Bergamaschi et al^5^ and re-analysed here for comparison with proteomic data

(C) Details of donors analysed by flow cytometry panels in Bergamaschi et al^5^ and re-analysed here to determine sample neutrophil contamination

(D) Details of donors used to validate and phenotype marker upregulation by flow cytometry

**Table S2**

Enrichment of functional pathways in clusters of cellular proteins upregulated during COVID. DAVID enrichment terms and corresponding Benjamini-Hochberg-corrected p-values are shown for each cluster in **Fig. 2B**.

**Table S3**

(A) Interactive searchable spreadsheet containing all data and statistics from WCL, PM and RNAseq analyses

(B) Proteomic data from all WCL analyses

(C) Proteomic data from WCL analyses for proteins quantified across all three WCL experiments

(D) Results of statistical tests comparing relative abundance of each protein quantified in WCL analyses.

(E) Proteomic data from second PM analysis

(F) Proteomic data from all PM analyses

(G) Results of statistical tests comparing relative abundance of each protein quantified in second PM analysis.

(H) Transcriptomic data from all donors generated in Bergamaschi et al^5^ at day 0 timepoint. Data expressed as Log_2_(RPKM).

(I) Transcriptomic data from donors also analysed in proteomic analyses, generated in Bergamaschi et al^5^ at day 0 timepoint. Data expressed as Log_2_(RPKM).

